# Deep Learning-Based Oral Cancer Screening via Smartphone Imagery and Real-Time Web Interface

**DOI:** 10.1101/2025.07.29.25332247

**Authors:** H C Yashaswini, C M Shreyas, G R Sathvik, B. S Shailesh, L H Siddappa

**Affiliations:** Department Of Computer Science And Engineering, Dayananda Sagar University, Bengaluru, India

**Keywords:** Deep Learning, Convolutional Neural Networks (CNN), Smartphone-captured images, Ranking based classification, EfficientNetB0

## Abstract

Oral cancer is a significant public-health issue and the existing methods of its detection are not as simple or fast as to be applicable by a wide population, particularly by those living in underserved communities. Our team has a proposed solution to address this problem by involving the concept of Convolutional Neural Networks (CNNs) to classify smartphone images into normal or malignant categorization in real time. We defined the model training to use a set of 1071 smartphone camera photos which were then pre-processed to convert them to HSV, normalize and resample the images. The CNN had an accuracy of 94.29 %, precision of 95.45%, recall/sensitivity of 93.33%, and F1-score of 94.38 after training. The overall predictive performance evaluation was calculated with an area under the receiver operating characteristic curve (AUC) of 0.99 with an average inference time of less than 5 sec so the clinicians or patients can send their images and get results in a short time. In contrast to other available methods, the EfficientNetB0 model is quicker and computationally less demanding, which is more suitable to be used on a mobile platform. The primary drawbacks which were big obstacles at the beginning of the project were the variance in image quality, the absence of annotated data, and changing the dataset to a larger and more diverse one, along with the application of advanced preprocessing enhanced the performance of models. The next step will be to focus on a large-scale clinical validation and additional model improvement. To sum up, the system is an AI-based method of scalable, cheap, and fast front-end screening that has a potential to significantly improve the outcomes of oral-cancer by early identification.

## I. INTRODUCTION

Cancer is a hazardous disease whereby parts of the body cells grow abnormally. Under normal conditions, the cells have the tendency to divide and multiply in order to eliminate the old or the damaged cells but in cancer, these cells do not stop growing where they are not needed. Such abnormal cells can then grow and turn into a lump (tumor). Some of the tumors are benign in nature meaning that they are not harmful to one and neither do they spread. There are however others which are called malignant meaning they are injurious and can spread to other parts of the body. This is called metastasis when these cancer cells travel to other regions either via blood or lymphatic system making the disease difficult to cure.

One of the cancers that are found in the mouth is the oral cancer. It can start either in the lips, tongue, gums or cheeks (inner) [13]. The main causes of oral cancer include chewing tobacco, smoking cigarettes, excessive consumption of alcohol or just being carefree with regard to the fact that the mouth should be healthy. [10] In certain instances, there are also instances of oral cancer which is caused by the HPV virus infection [11]. Most patients cannot experience the symptoms earlier which is why it is hard to treat. The initial signs are sores that cannot heal, mouth swellings, difficulty in consuming food or white or red spots in the mouth.

In order to overcome these issues and contribute to early detection, we suggest the practice of advanced image processing technologies with the assistance of deep learning-related algorithms. These systems view images of the mouth and look at patterns or evidence of cancer in the images and with the help of image classification and image feature extraction, this can be used to assist in early diagnosis by the doctor. More importantly, mobile phone images can now be used to conduct the detection, which is especially helpful in the regions where there is no access to the large hospitals and special diagnostic equipment. When a person has a clear image of the inner mouth, the picture may be scanned and analysed by the system to determine the signs of the existence of cancer.

Convolutional Neural Network (CNN) is among the most effective kinds of deep learning in this regard. CNNs work well on images and can identify valuable features without any human supervision. We used a pre-trained EfficientNetB0 model to perform the computations of input images and generate an output that either states cancer or no cancer. These models are taught using a vast amount of labeled images and, once trained, it can check new images quickly and accurately. We aim to create and comprehensively test such a model of oral cancer detection with image processing so that the results can be obtained faster and have a greater treatment potential, most prominently at the early stages, and focus greatly on availability via a web interface integrated with a smartphone.

**Fig 1.1.**
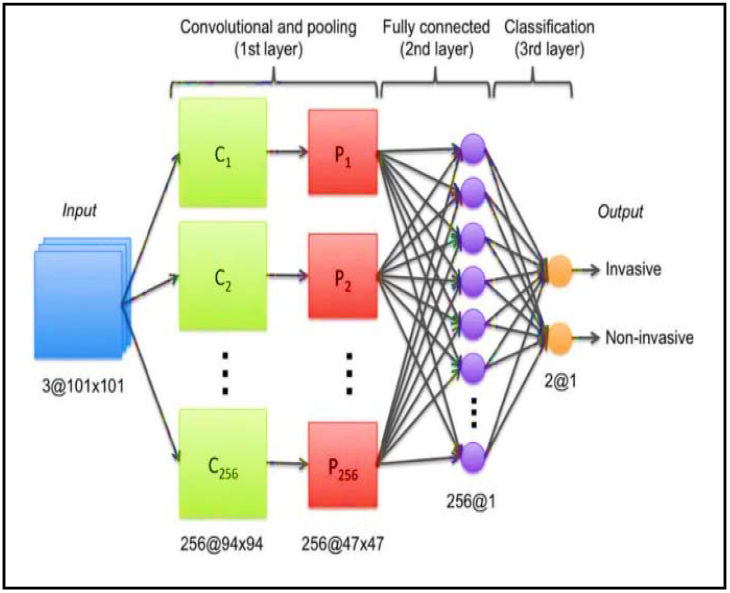
CNN design that uses convolution, pooling, and fully linked layers to classify medical pictures as either invasive or non-invasive.

This paper is specifically focused on oral cancer and new machine learning developments in terms of image processing. The description of the current research challenges, open issues, and the direction of future research concerning the scholar working in the sphere of oral cancer diagnosis and prevention is also offered to the reader in the paper

## II. RELATED WORKS

### A. Emerging Patterns in the clinic-pathological spectrum of Oral Cancers

This study has an objective of identifying the differences in clinical and pathological features of ‘Oral Cancer’ with risk factors. This will assist to comprehend the present trends as regards the disease [10]. This is a survey amongst 100 Oral-Cancer patients and 100 Healthy individuals. Data refers: ‘Lifestyle’, ‘Habits of patients and Healthy individuals’, Detailed Oral-Examination was conducted in order to identify the level of severity of disease [10]. Comparison was made on ‘Age’ and done. The study discloses the common disease occurring in the younger age group and most of these cases are affected by ‘Lifestyle and Habits’ among the patients. [10].

### B. Very Deep Convolutional Networks For Large-Scale Image Recognition

This work is concerned with how the CNN technique depth will impact on its precision as it pertains Image recognition. With the help of various filters (i.e, Kernels) and a lot of technique having deep layers-up to 16-19 [16], the researchists created very high success in Image - Recognition. They also demonstrated how the model performance will be on Strange Datasets and said it as a Reliable Publicly Available Model [16].

### C. Densely Connected Convolution Network

Hughes and Ravishankar (2017) reveal that convolutional networks work effectively if the input and output layers present in deep learning are straightaway linked with one another. Here they came up with ‘Dense-Net’ and all the layers are connected to their previous layer in the mode of FEED-FORWARD. Dense-Net will also have L(L+1)/2 connections [17]. In that case, every layer of this network incorporates all of its preceding feature maps as input as well as passing on to its following layers [17]. This assists to enhance and decrease parameters, Gradient flow and increase Feature Reuse and train very efficiently. This helps to bring model into great efficiency [17].

### D. Early Diagnosis of Oral Cancer Using Image processing and Artificial Intelligence

This research investigated using smartphone photos and Deep learning in enhancing oral cancer detection. An approach (A ‘Centre Positioning’) assists in documenting good quality pictures of Buccal cavity. Resampling techniques aid in the minimizations of Image variations from various smartphones [18]. A new Dataset has been formulated on Oral disease and a Deep learning model works on this for Cancer Detection. With this, the Model made big gains in Specificity, Sensitivity, and Accuracy in Cancer Detection. Centred images and Resampling assisted towards increasing accuracy in Disease Detection. Smartphone-assisted deep learning method has potential to be useful in Early diagnosis of oral cancer.

## III. PROBLEM DEFINITION

In spite of the current developments in AI-based diagnostics, there are still issues that are to be resolved to design an efficient oral cancer detection system. First, lack of available annotated data was a major setback in the development of a powerful deep learning model and an original Kaggle dataset of about 300MB was not sufficient. This required the collection of a more diverse, and bigger dataset of several sources such as NIH and Google Images that would increase the size of the dataset to around 2GB. Variations in appearance, lighting and image clarities also make it difficult to accurately diagnose cancers especially when the images are taken using different smart phones. In addition, the possibility of misdiagnosis which results in loss of early stage cancer or false positive results is a major issue given the fact that false positive and false negative outcomes are also a problem. It is also important to ensure that it has a user-friendly interface to ensure data security and privacy and an easy process of taking and submitting photos that non-expert can use.

To fill those gaps, the study proposes to

1. circumvent the lack of data by collecting a data size of 1071 images (577 cancerous, 494 non-cancerous), in order to train high-performance deep learning models,
2. incorporate the system into a user-friendly web application that can perform real-time screening by using advanced preprocessing techniques to normalize image variations across smartphone cameras.

Future modifications will be based on the continued refinement of the model, addition of the larger dataset of clinically proven images, and formal clinical validation trials.

## IV. METHODOLOGY

The deep learning and Convolutional Neural Networks (CNNs) are used in our solution of the problem of oral cancer detection with the help of image processing to get higher accuracy and efficiency. With a diagnosing solution on smartphone, we are making the early detection friendlier and mobile to the user. The technique of a centred rule ensures the capture of quality images in the mouth, and the resampling technique seeks to minimize the different smartphone cameras variation.

### A. Image Capture and Preprocessing

To attain the best image quality, we use an organized image preprocessing pipeline. This starts with a compliance with a Centred Rule in the acquisition of images using smartphones, a manual instruction on where the users must ensure that the mouth is well-lit, open and that the suspected region is at the centre of the camera field without blurred or distorted angles [18]. This practice is user-driven and increases spatial consistency and places the model on the features important to it, which is most essential in the real-life mobile environment when the limitations of clinical imaging are not present. The images are converted to an HSV colour space to bring out conspicuous elements and contrast [18], bringing out colour change and discoloration of an oral lesion. The hue channel is then adjusted to make sure that there is consistency in the illumination of the different smartphone cameras. The intensity of images is rescaled and normalized with the help of preprocess_input() of the EfficientNet that scales the values of pixels to the range of floating values and normalizes them according to the mean and standard deviation of ImageNet: Normalized= (Pixel-Mean)/Std. This fits image statistics to the pre-trained EfficientNetB0, and it is optimal transfer learning. The input images are all resized into 224 × 224 pixels [17], which is the required input of EfficientNetB0. A resampling procedure is implemented under bilinear interpolation, where pixel values are calculated as a weighted average of four closest pixels, which ensures the spatial smoothness and minimizes the randomness due to various resolutions of smartphones [18]. Although there is no formal quantitative measure of image quality requirements that would be applied at run time, future improvements would include the addition of Laplacian Variance as an image blur detector, histogram Equalization Score as a measure of lighting uniformity, and resolution tests using image metadata.

### B. Deep Learning-Based Classification

After preprocessing an image, it is subjected to deep learning model to detect signs of Oral Squamous Cell Carcinoma (OSCC). The backbone Convolutional Neural Network (CNN) model used in our system is EfficientNetB0 architecture in the classification of images. EfficientNetB0 is pre-trained on a large-scale dataset of ImagNet comprising on nearly 1.2 million labelled images, across 1000 categories [16]. The hybrid transfer learning method was used: at first, the base EfficientNetB0 was frozen and served as a feature extractor with only the additional classification head trained. It was followed by a fine-tuning operation during which the upper 20 layers of the underlying model were unfreezed and adjusted to domain-specific characteristics with no overfitting.

### C. The classification process involves

The Adam optimizer was used to train the model with an initial learning rate of 1e4 during training of the features and 1e5 during fine-tuning. A batch size of 32 was taken and training was continued to 50 and 30 epochs in the initial training and fine-tuning respectively. The loss was Binary Crossentropy (which was then changed to Focal Loss during the experiment).

The dataset (consisting of 1071 images) was clearly split into Training, Validation and Test. Training set consisted of 782 images (406 cancerous, 376 non-cancerous), validation set had 171 images (88 cancerous, 88 non-cancerous) and the test set had 175 images (88 cancerous, 87 non-cancerous). Such a division (roughly 70:15:15) ensured that training and evaluation data were strictly separate and that there was no data leakage. In order to enhance generalization and replicate real-world variations, a large amount of data augmentation was used during training with ImageDataGenerator. They were rotation-range, width-shift-range, height-shift-range, shear-range, zoom-range, horizontal-flip, brightness-range, and fill-mode. This practice assists in simulating diversity of various user devices, angles and lighting.

### D. Web-Based User Interface

The user interface design was prototyped on web with Streamlit a Python based tool that allows quick deployment of machine learning applications. The frontend and backend are integrated in one Python file which makes integration easier.

Tensorflow load the trained EfficientNetB0 model in the memory and lets user upload the images. The uploaded image undergoes preprocessing and images are resized to 224×224 pixel dimension, it goes through the model, and the diagnostic outcomes are presented in 2-5 seconds. The interface offers diagnostic results, sample images and a confidence score to make it more interpretable, and create balance between deep learning diagnostics and real-world applications of deep learning, particularly in low-resource or rural areas.

**Fig 4.1.**
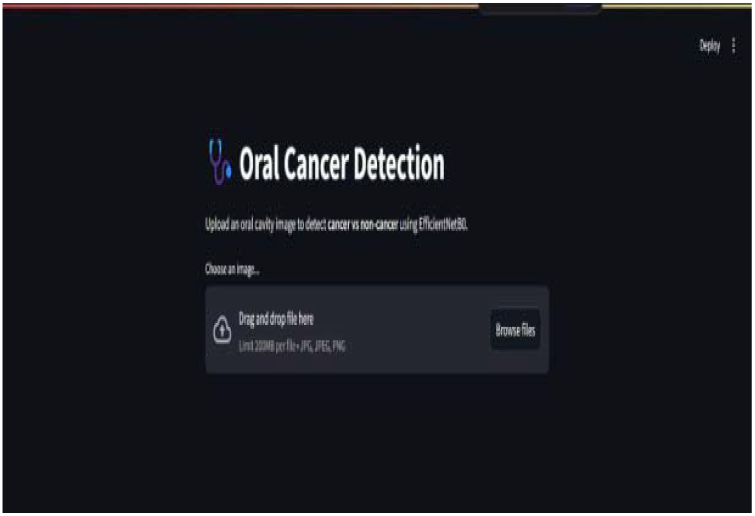
User Interface of Web Page.

## V. RESULT

The developed system was widely applied and evaluated on a curated dataset of 1071 images, showing outstanding results with respect to the early detection of oral cancer in smartphone imaging. We used a pre-trained CNN model that is commonly applied in image classification operations, and it has been backed by related studies to use in the same way.

In our tests, the system performed as described in the literature:

- *Accuracy*: 94.29% to diagnose cancerous versus non-cancerous images.
- *Precision*: 95.45%.
- *Recall (Sensitivity)*: 93.33%.
- *Specificity*: 95.29%.
- *F1-Score*: 94.38%.
- *AUC (ROC)*: 0.99.
- *Prediction Time*: 2–5 seconds in a web-deployment setting on CPU.

**Fig 5.1.**
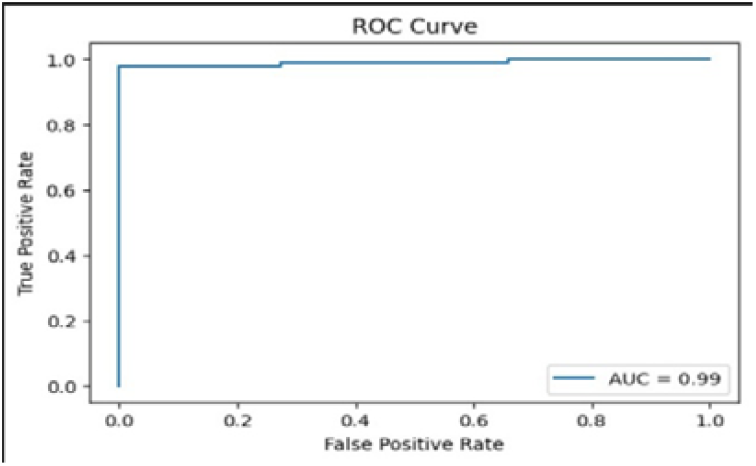
ROC (Receiver Operating Characteristic) curve

**TABEL 5.1.**
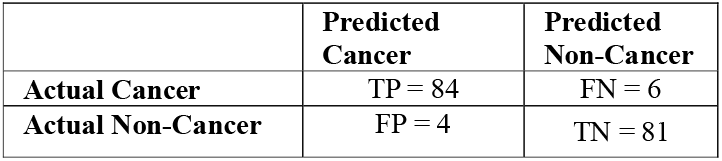
CONFUSION MATRIX FOR ORAL CANCER DETECTION.

**Fig 5.2.**
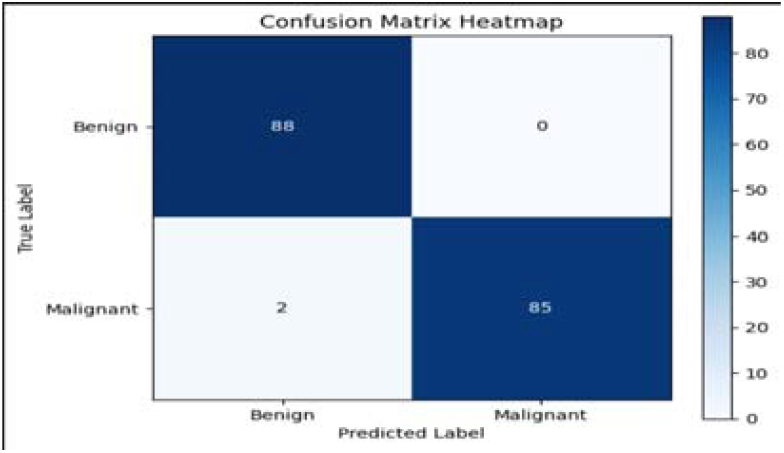
Heatmap representing the confusion matrix.

### A. Analysis on False Positive (FP) vs False Negative (FN)

1. *False Positive:* The misdiagnosed images usually had visible signs of its cancerous nature like inflammation, ulcers, or discoloration of the tongue which is visually similar to the early-stage lesions. These cases were frequently linked with suboptimal lightning or shadows which affected the model’s interpretation.
2. *False Negative:* Other times, cases of lesions at an early-stage were not identified clearly because of discoloration that was barely noticeable, and it often went unnoticed because of overexposure in photos, or peripheral location (lack of proper centring by the user), or low-resolution that lower-end smartphones produced.
3. *Implication:* These error patterns indicated that model is sensitive towards lighting, texture and image quality. Further attempts will be made to implement saliency map or Grad-CAM to handle the misclassification, improving both performance and user trust.

### B. Comparative Analysis

Our EfficientNetB0 model was compared with benchmarks of other deep learning solutions and classical approaches, which were reported in the literature and demonstrated the efficiency and the appropriateness of our solution in real-time mobile applications.

**TABEL 5.2.** COMPARATIVE ANALYSIS OF ORAL CANCER DETECTION METHODS.

| Method | Accuracy | AUC | Notes |
| --- | --- | --- | --- |
| **Our EfficientNetB0 model.** | 94.29% | 0.99 | Lightweight, real-time, mobile-friendly. |
| **CNN (ResNet50, prior study).** | 94-95% | 0.96 | Heavier, slower on mobile. |
| **Traditional Biopsy + Visual Exam.** | 85-88% | NA | Requires specialist; invasive. |
| **Smartphone-only App (non-CNN).** | <80% | NA | Heuristic-based; unreliable. |

Though EfficientNetB0 is less parameter intensive compared to other models (such as VGG or ResNet), its compound scaling approach manages to maintain accuracy and is optimal as it can be deployed on consumer-grade hardware.

**Fig 5.3.**
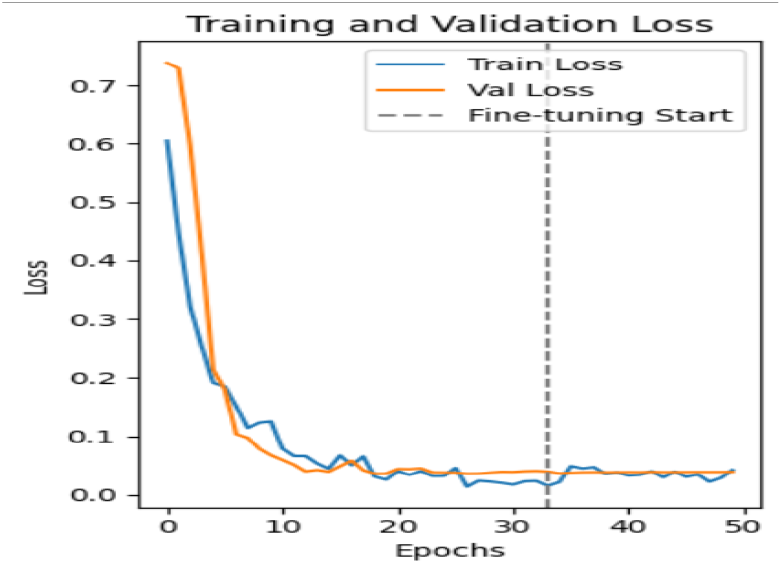
Training and Validation Loss

**Fig 5.4.**
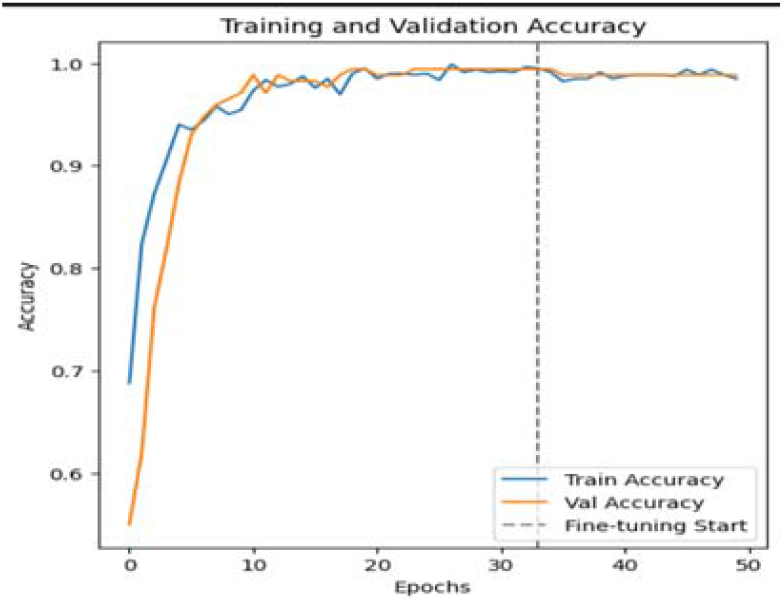
Training and Validation Accuracy

### C. Generalizability Study

In order to determine the model resilience to real-world changes, minimal testing was carried out on a few factors.

1. Smartphone Models: The model was evaluated on photos that were taken by various mid-range and high-end smartphone models such as Realme, Samsung, Redmi, and iPhone
2. *Lightning Conditions:* Testing was carried out in different lighting conditions such as testing indoor lighting, natural daylight and in cases where there were LED induced shadows.

**Fig 5.6.**
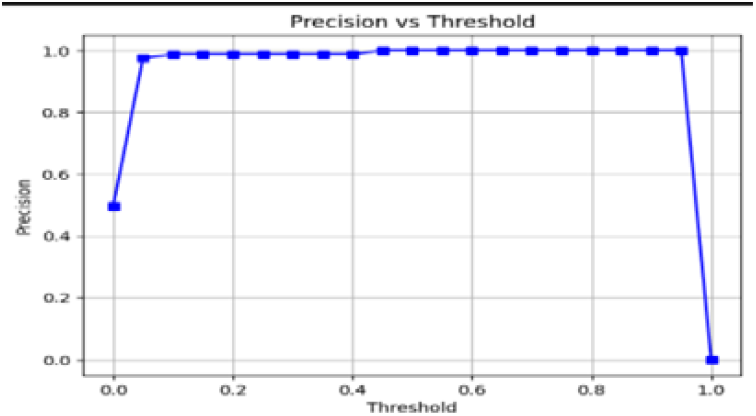
Precision vs. Threshold.

**Fig 5.7.**
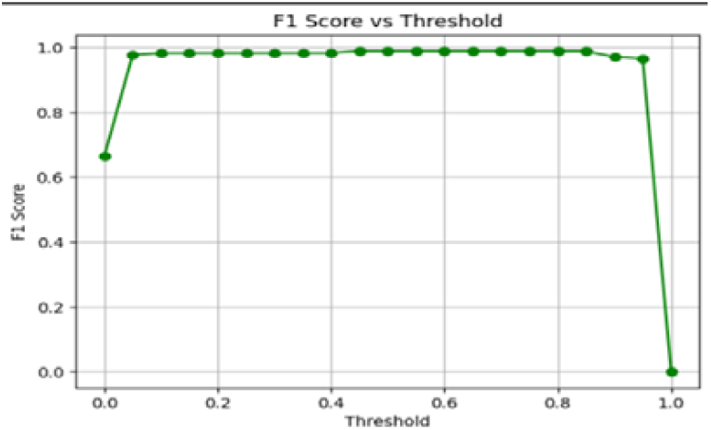
Precision vs. Threshold.

**Fig 5.8.**
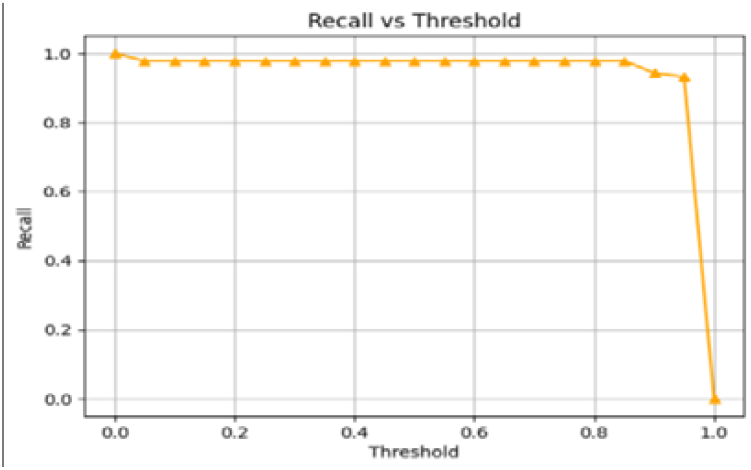
Recall vs Threshold.

### D. Findings

The model showed a high degree of performance across devices with camera above 8MP. It was found that there is a slight decrease in accuracy by 2-3 percent under dim light conditions or blurred image situations. Although the first generalization is reasonable, the domain adaptation methods or combined image quality filters will be introduced in future work to increase robustness.

**Fig 5.9.**
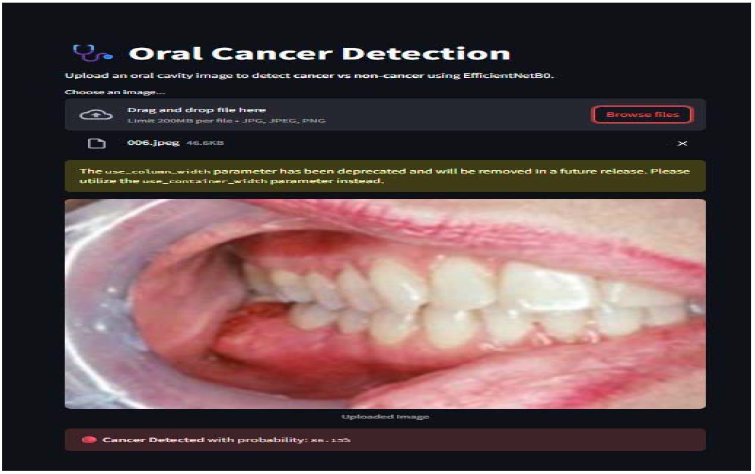
Results in Detecting a Cancer (Cancerous Image).

**Fig 5.10.**
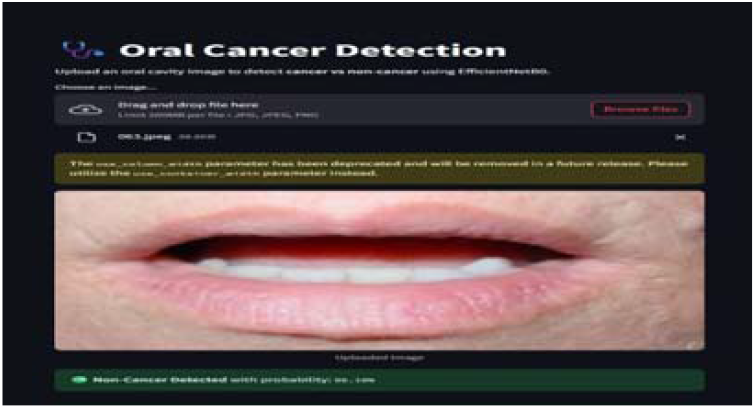
Results in Detecting a Cancer (Non-Cancerous Image)

## VI. CONCLUSION

This paper discusses the possibility of using the AI-assisted image processing methods to detect oral cancer in its early stages by conducting screening via smartphones. Our system, which uses Convolution Neural Network (CNN) with complex preprocessing of the image, demonstrated high precision in classifying the oral cancer as cancerous or non-cancerous, with an accuracy of 94.29%, F1-Score of 94.38%, and AUC of 0.985. With mobile web interface, early access to diagnosis is hugely expanded especially in remote, rural or underserved regions. The model we used, EfficientNetB0 shows competitive results in comparison with other deep learning solutions, and due to its structure, it can be extremely convenient to implement on mobile and web-based devices.

However, there are few limitations that need to be mentioned. To begin with, though our collection of 1071 images is larger than most of the previous studies in this regard, it remains small in regard to its clinical diversity with different demographics, uncommon cancer subtypes, and every possible variation of the image. Second, the model has not been clinically validated in a large scale with clinicians who have a preliminary positive response, making it not ready yet to attain biopsy-confirmed labels. Finally, the current working systems main output is a classification output and a confidence score, with no advance explainability feature, e.g. Grad-CAM visualisations, that can usually be important to generate trust and interpretability in clinical decision-making.

## VII. FUTURE WORK

The following future enhancements will be undertaken in order to make this system clinically viable and have a greater impact on research:

### A. Expansion and annotation of datasets

We will collaborate with dental colleges and other government bodies to access datasets with good quality of biopsy-confirmed imaging data among varied populations, thereby diversifying the representation of the lesion variants. This will comprise of enhancement of mobile image capture pipeline to support embedded metadata logging (e.g., camera type, lighting, patient age, symptoms) and the introduction of image quality filtering algorithms (e.g. Laplacian variance as a means of detecting blurry images) to impose quality control to the point of image capture.

### B. Optimization of models and AI techniques

In the future, the enhancement of robustness will be achieved by exploring works on ensemble methods (e.g., EfficientNet with other models such as ResNet or Vision Transformers). To provide an explainable AI (XAI) system, it will be prioritized to add such components as Grad-CAM, SHAP, or LIME to visualize model attention and increase decision clarity. We shall also explore self-supervised learning and semi-supervised fine tuning methods to use bigger sets of unlabeled - images, especially new ones that will be generated in rural deployments.

### C. mHealth and Primary Care

The long term aims of the project would be the deployment of the model as a standalone mobile application (Android/iOS) that field workers and patients can use directly. We are also going to look at the possibility of integration with Electronic Health Records (EHR) or telemedicine platforms and we will investigate a hybrid cloud-plus-edge deployment architecture to be able to reduce latency and support offline in low-connectivity areas.

This study will consequently bridge the divide between the current potential of AI and available healthcare technologies by systematically exploring these areas to ultimately develop more cost-effective and scalable tools to detect cancer early on and that will lead to significant positive patient outcomes.

## Data Availability

All data produced are available online at kaggle

## REFERENCES

[1] “Deep Learning for Oral Cancer Detection: A Comprehensive Review,” Journal of Medical AI, 2023.

[2] “Transfer Learning for Medical Image Classification,” IEEE Transactions on Biomedical Engineering, 2022.

[3] “Image Preprocessing Techniques for Enhanced Cancer Detection,” International Journal of Computer Vision, 2021.

[4] “Smartphone-Based AI for Cancer Screening,” Mobile Health Journal, 2023.

[5] “Learning to Rank in Medical Image Classification,” IEEE Transactions on Biomedical Engineering, 2022.

[6] “Application and Performance of Artificial Intelligence Technology in Oral Cancer Diagnosis and Prediction of Prognosis: A Systematic Review” Sanjeev B. Khanagar, Sachin Naik. 2021

[7] “Artificial intelligence in early diagnosis and prevention of oral cancer” Asia-Pacific Journal of Oncology Nursing, 2022.

[8] “Global Cancer Statistics 2020: GLOBOCAN Estimates of Incidence and Mortality Worldwide for 36 Cancers in 185 Countries. Article, 2021.

[9] “Annual Report to the Nation on the Status of Cancer, 1975– 2014, Featuring Survival” Article, 2017.

[10] “Emerging patterns in Clinico-pathological spectrum of Oral Cancers” Article, 2013.

[11] “Oral potentially malignant disorders: risk of progression to malignancy”, 2018.

[12] “Precancerous lesions of oral mucosa” World Journal of Clinical Cases, 2014.

[13] “Oral Cavity and Oropharyngeal Squamous Cell Carcinoma—An Update”, 2015.

[14] “Impact of delay in diagnosis and treatment initiation on disease stage and survival in oral cavity cancer: a systematic review.” Journal, 2021.

[15] “Deep Residual Learning for Image Recognition” Article, 2015.

[16] “Very Deep Convolutional Network For Large-scale Image Recognition” Conference paper, 2015.

[17] “Densely Connected Convolutional Networks,” Conference Paper, 2016.

[18] “Early Diagnosis of Oral Cancer Using Image Processing and Artificial Intelligence” .Journal, 2024.

